# Reward-related activation of fronto-striatal regions scaled negatively with C-reactive protein but showed no association with anhedonia in depression

**DOI:** 10.1101/2022.10.05.22280729

**Authors:** Athina R. Aruldass, Manfred G. Kitzbichler, Tsen Vei Lim, Wellcome Trust Consortium for Neuroimmunology of Mood Disorders and Alzheimer’s Disease (NIMA), Jonathan Cavanagh, Phil Cowen, Carmine M. Pariante, Edward T. Bullmore, Neil A. Harrison

## Abstract

Depression is characterized by divergent changes in positive and negative affect. Emerging roles of inflammation in depression portend avenues for novel immunomodulator-based monotherapy, targeting mechanistically distinct symptoms such as anhedonia and pessimism. To investigate fundamental links between these divergent affective components and inflammation, we used a probabilistic reinforcement-learning fMRI paradigm, testing for evidence of hyposensitivity to reward, and hypersensitivity to punishment in low-inflammation depression cases (loCRP depression; CRP ≤ mg/L; N=48), high-inflammation depression cases (hiCRP depression; CRP > 3mg/L; N=31), and healthy controls (HC; CRP ≤ mg/L; N=45). We aimed to (i) determine whether depression cases with high and low inflammation showed aberrant neural activation to monetary gains and losses compared to controls; (ii) examine if these alterations correlated with a continuous measure of C-reactive protein (CRP) in depression, (iii) test if neuroimaging responses to rewards and punishments scaled with indices of anhedonia and pessimism derived from behavioral instruments in depression. Voxel-wise activation was observed in key brain regions sensitive to monetary reward (ventromedial prefrontal cortex, vmPFC; nucleus accumbens, NAc) and punishment (insula) outcomes across all three groups. However, there was no significant difference in activation between groups. Within depression cases, increasing CRP scaled negatively with activation in the right vmPFC and left NAc but not insula cortex. However, there was no significant association between regional activation and severity of anhedonia or pessimism. Our results support the previously reported association between CRP and striatal reward reactivity in depression but do not extend this to processing of negatively valenced information.

## Introduction

Cognitive vulnerability (CV) is central to many prominent theories of the pathogenesis of major depressive disorder (MDD). This includes Beck’s pioneering theories on negative schemata, i.e., dysfunctional internal beliefs or pernicious self-representations such as being highly self-critical (1); Abramson’s hopelessness theory (2); and Hoeksema’s response styles theory focused on maladaptive rumination or brooding (3). Collectively, these theories highlight the insidious link between increased negative affect and depression. Supporting this, fMRI studies over the past decade have demonstrated maladaptive response to negatively valenced events such as monetary loss or social defeat, hypersensitivity to punishment or larger error-related negativities reflected by increased insular activity in MDD cases (4–7). However, the biological processes that drive processing of negative emotional states in the insula remain to be delineated. Consequently, interventional strategies for managing cognitive vulnerabilities in depression broadly focus on cognitive behavioral therapy approaches geared towards “undoing” maladaptive information processing, rather than pharmacological or other more biologically-oriented interventions (8).

However, such depressogenic cognitive schemata are also thought to play a role in precipitating more ‘biologically embedded’ features of depression such as anhedonia which are often targeted pharmacologically. For example, anhedonia, a core diagnostic symptom of MDD, alongside low mood, has been linked to functional impairments in several subdomains of reward processing (9) with meta-analyses reporting reduced striatal activation during reward-related processing in depression (10–12). In particular, using experimentally controlled rewards, patients with MDD exhibit ventral striatal (specifically nucleus accumbens: NAc) hypoactivation (13–17), and impaired medial prefrontal cortex (mPFC) (16,18–20) activation compared to healthy controls. Despite this fMRI evidence for a biological substrate of aberrant reward processing, anhedonia is often a particularly difficult symptom to treat as first-line pharmacotherapies, such as selective serotonin-reuptake inhibitors (SSRIs), selective noradrenaline-reuptake inhibitors (SNRIs), and tricyclic antidepressants (TCAs), do not effectively target the motivational and hedonic deficits in depression, which are largely encoded by dopamine (DA) (21–23).

Fundamentally, the dichotomy of negative and positive emotion processing abnormalities in depression illuminates the dissociable neurobiological foundations of MDD: namely, (i) up-regulation of a negative valence system and (ii) down-regulation of a positive valence system (24,25). As such, the ideal pharmacotherapy would target both negative affect and positive affect systems i.e. lowering negative affect and increasing positive affect simultaneously (24–27).

Recently there has been growing interest in the pathogenic role of chronic low-grade peripheral inflammation in a subset of patients with depression (28–30). This association between immune mechanisms and depressive psychopathology may offer novel avenues for pharmacotherapy, such as immunomodulatory drugs (31,32), that could potentially target both aberrant negative and positive affect processing systems. Indeed, studies have shown robust evidence for a link between peripheral inflammation and depressive symptoms, particularly anhedonia (33) and, to a lesser extent, negative affect (34). Most prior investigations were experimental models of inflammation-linked depression, where healthy humans (or animals) were experimentally treated with a pro-inflammatory stimulus to induce sickness behaviors that show striking similarities to depression, including symptoms such as malaise, anergia, low mood and anhedonia. In human models, multiple fMRI studies have demonstrated reorientation to reward- and punishment-related stimuli following pro-inflammatory challenges (17,35).

Noteably, despite using distinct activation paradigms two independent studies recently reported convergent hypersensitivity or increased activation within brain regions of the ‘neural alarm system’ (36,37) i.e. the anterior insula (34) and amygdala (38), in response to negatively valenced stimuli of monetary penalty or receipt of negative social feedback, respectively following pro-inflammatory challenge. Inflammation-related changes in brain activation related to positive affect or positively valenced stimuli have been more extensively investigated. For example, endotoxin challenge resulted in hypoactivation of the ventral striatum during hedonic reward anticipation in a monetary incentive delay (MID) task (39).

Corroborating this observation, Capuron et al. (2012) (40) reported a similar reduction in bilateral ventral striatal activity for winning versus losing outcomes in a gambling task administered to hepatitis C-seropositive (HCV+) patients receiving IFNα therapy compared to HCV+ patients awaiting therapy. Notably, inflammation-related changes in task-activated reward processing were correlated with observer ratings of anhedonia.

Supporting this perspective, observational fMRI studies of inflammation-linked depression – predominantly resting-state fMRI reports – have shown evidence for similar alterations to functional connectivity of mesolimbic or reward circuitry in depression in the context of chronic low-grade peripheral inflammation. In particular, a seminal observational case-only study demonstrated reduced fronto-striatal (ventromedial PFC – ventral striatum) connectivity with increasing CRP, which in turn correlated positively with inter-individual differences in anhedonia (33). Analogous findings have been reported in case-control studies that have shown reduced functional connectivity between affective brain regions such as the prefrontal, cingulate and insular cortices in inflammation-related depression (41,42). However, such resting-state fMRI studies are inevitably lacking in experimentally controlled cognitive or behavioural conditions to support psychopathological interpretation of case-control differences in brain function.

Here, we investigated abnormalities of reward and punishment processing in inflammation-associated depression using a probabilistic reinforcement learning fMRI task. We aimed to (i) ascertain whether depression cases with high and low levels of peripheral inflammation showed aberrant neural activation to monetary gains and losses, compared to healthy controls (HCs); (ii) examine if these alterations correlated with a continuous measure of CRP in depression cases; and (iii) test if the neuroimaging data correlated with behavioral indices of anhedonia and pessimism in depression cases. We hypothesized that, in depression cases, receipt of a monetary gain or reward would result in hypoactivation of reward-related brain regions, e.g., ventral striatum and mPFC, and scale negatively with blood concentration of CRP. In contrast, we hypothesized that receipt of a monetary loss or punishment would result in hyperactivation of punishment-related brain region(s), e.g. anterior insula, and scale positively with CRP in depression cases. Further, we also hypothesized that depression cases with (i) hypoactivation of reward-related brain regions would exhibit higher odds of more severe anhedonic attitudes and (ii) hyperactivation of punishment-related brain regions would demonstrate higher odds of more severe negative attitudes.

## Methods & Materials

### Participants

Biomarkers for Depression (BioDep) was an observational case-control study conducted as part of the Wellcome Trust Neuroimmunology of Mood Disorders and Alzheimer’s disease (NIMA) Consortium. All procedures were approved by an independent national research ethics service (NRES) committee (NRES: East of England, Cambridge Central, UK; Reference: 15/EE/0092) and all participants provided written informed consent.

All participants satisfied inclusion criteria, e.g. aged 25-50 years, and exclusion criteria, e.g. major medical inflammatory disorder or immuno-modulatory medication, as reported in our earlier investigation (41). All depression cases screened positive for current depressive symptoms on the Structured Clinical Interview for DSM-5 Depressive Disorders (SCID) and had total Hamilton Rating Scale for Depression (HAMD-17) score > 13, as reported previously (41).

After initial telephone screening, potentially eligible participants attended an eligibility assessment, including blood sampling for CRP assay, at one of 5 UK recruitment centers (Brighton, Cambridge, Glasgow, King’s College London (KCL), or Oxford). Eligible participants next attended one of 3 UK assessment centers (Cambridge, KCL or Oxford) for venous blood sampling, clinical assessment, and MRI scanning. All blood samples were scheduled for the same time of day (8-10am). Blood plasma collection and high-sensitivity CRP assaying are detailed in the **Supplementary Material (Sections1-2)**. Depression cases were then stratified by blood CRP level: low CRP depression cases had CRP ≤ mg/L, high CRP depression cases had CRP > 3mg/L. Three high CRP depression cases with CRP > 10mg/L (10.2 – 11.4 mg/L) were retained for analysis as there was no clinical evidence for infection or other exclusionary medical disorders. All HCs had CRP ≤ mg/L.

### fMRI reward and punishment paradigm

Participants completed a probabilistic reinforcement learning paradigm (**Figure 1**) (34,43,44) where the ultimate goal was to maximize rewarding monetary gains or winnings, and minimize punitive monetary losses. The task comprised 3 trial types, i.e., ‘Gain’ condition (reward), ‘Loss’ condition (punishment) and ‘Look’ condition (neutral), with 24 trials of each type randomly interspersed in a single run (72 trials in total) (**Figure 1**). On each trial, participants were required to choose between two visual stimuli (or cues) displayed on a screen for 4s, before making a selection, i.e. right-sided stimulus by triggering button press (GO response), and the left-sided stimulus by withholding response (NO-GO response). The selection (choice made) was then circled in red and the outcome (or feedback) displayed on the screen after a 4-second delay. Outcomes for Gain, Loss and Look trials were gain +£1/ nil, loss -£1/nil and nil/nil respectively (with £1 represented by a pound coin, and nil feedback by a ‘nothing’ sign of equal size as a pound coin, both shown on a visual display screen for 4s). The feedback was probabilistic in that between gain and loss conditions, there was a fixed probabilistic 80:20 contingency of positive feedback. At the end of the scanning session, participants had the option to find out their total earnings. However, they were not paid extra money (on top of the standard reimbursement for BioDep participants). The task was coded using MATLAB. Participants performed a practice trial of the task initially on a computer/laptop, before performing it a second time in the scanner.

**Figure 1.**
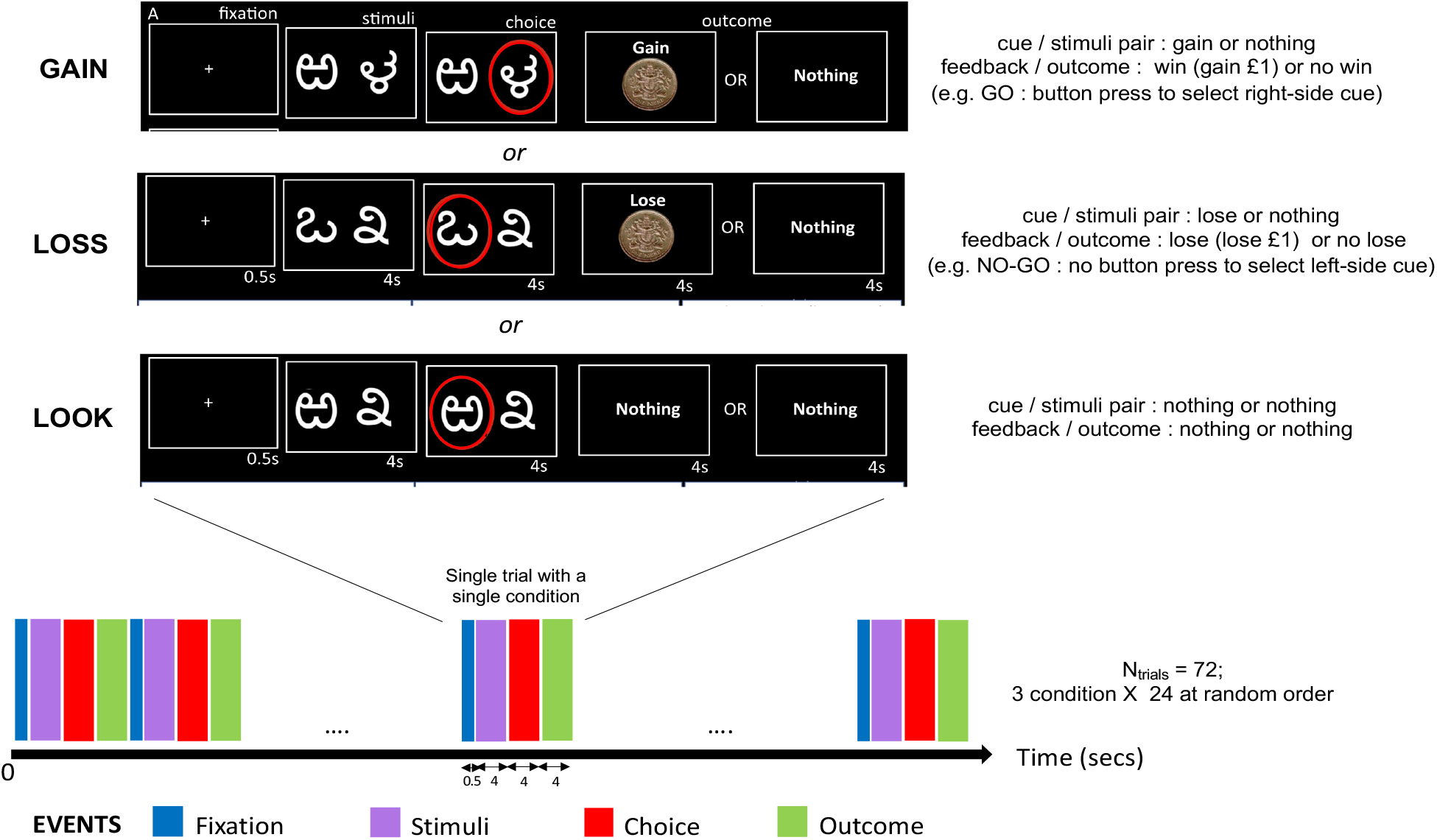
Probabilistic reinforcement learning paradigm. The task paradigm was a version of the probabilistic selection stimulus task, with an event-related design. Participants performed the task over a single run, with a total of 72 trials (24 trials per condition, i.e., GAIN, LOSS, LOOK) presented in random order. *Adapted from Harrison et al. (2016) Biological Psychiatry* (34).

### fMRI data analyses

Following preprocessing and estimation of motion regressors (see **Supplementary Material Section 1 and 4** for detailed fMRI data acquisition and preprocessing procedures), we performed first-level GLM analysis using AFNI’s *3dDeconvolve* function to identify regions responding to gain (re-ward) or loss (punishment) trials (**Figure S1**). Subjects with a high degree of head motion estimated by framewise displacement, FDmax > 1.3mm and/or FDrms > 0.3mm were excluded from the analysis. On this occasion, all participants passed quality control criteria for motion, none were subject to exclusion (N=0). Preprocessed timeseries for each subject were convolved with a gamma function modelling the canonical hemodynamic response function (HRF). The full regression model included a set of 11 orthogonal regressors of interest: presentations of cue or stimulus pair specific to each trial type, i.e., Gain, Loss or Look; response selection or button pressing event, i.e. GO versus NO-GO motor activity; gain outcomes (winning or +£1.00) versus non-gain outcomes (no win or ‘nothing’); loss (-£1.00) versus non-loss outcomes (‘nothing’); and ‘nothing’ outcomes for Look trials. We also included 12 regressors of non-interest or nuisance covariates comprising six regressors estimating rigid-body head motion, six regressors estimating derivatives of each of the motion parameters, and an additional 5 baseline regressors (*-polort* argument in *3dDeconvolve* set to 4).

Based on initial QC of computational and fMRI task data validity (see **Supplementary Material Section 9; Figure S2**), second-level hypothesis testing focused on changes in fMRI contrast (or “activation”) occurring during the feedback or outcome phase. The regression coefficients (contrast weights or betas) for each trial type in each participant were treated as the dependent variables in whole-brain voxel-wise *t*-tests using AFNI’s *3dttest++* function for the following contrasts: (i) Gain (win or +£1.00) versus non-Gain outcomes (no-win or ‘nothing’); and (ii) Loss (lose or -£1.00) versus non-Loss outcomes (no-lose or ‘nothing’). Group data were compared in two ways. First, we tested for whole-brain voxel-wise activation significantly different than zero in response to each trial type, within each group using one-sample *t*-test. Second, we performed four two-sample *t*-tests for between-group differences in trial-specific activation for HC versus all depression cases, HC versus low CRP depression cases, HC versus high CRP depression cases, and low versus high CRP depression cases. Statistical parametric maps were generated with an initial voxel-wise threshold at *PFDR* < 0.05 (q-value in AFNI GUI set to 0.05), and a criterion of at least 40 suprathreshold contiguous voxels as defined by the AFNI *3dClusterize* function. A more conservative threshold of *PFDR* < 0.001 (q-value in AFNI GUI set to 0.001) was applied where appropriate to increase spatial localization of activation signal and improve visualization.

### Confirmatory analyses using atlas-derived ROI masks

Based on second-level between-group analyses, and the previous literature on reward and punishment processing, the bilateral vmPFC (ventromedial prefrontal cortex), NAc (nucleus accumbens; component of the ventral striatum) and anterior insula were selected as ROIs for further analyses of correlations between task-related activation, CRP and behavioral indices using data from depression cases only. All ROIs were anatomically defined by standard prior atlases (**Table S1**) and were examined laterally i.e. left and right separately. For each ROI, the average contrast weights (beta estimates) from the GLM analyses of Gain (reward) and Loss (punishment) conditions were extracted using AFNI’s *3dmaskave* function.

### Assessment of reward- and punishment-related affective abnormalities

Reward- and punishment-related affective abnormalities in depression cases were assessed using the Beck’s Depression Inventory version 2 (BDI-II) (45). We examined subscales within the questionnaire encoding for two key affective components: (i) anhedonic attributes and (ii) negative attitudes. Four items were considered under anhedonic attributes: loss of pleasure (item #4), loss of interest (item #12), loss of energy (item #15), and loss of interest in sex (item #21), as utilized in earlier investigations of anhedonic subtypes (46,47). The negative attitudes component was weighted by pessimism (item #2), punishment feelings (item #6), self-dislike (item #7), and self-criticalness (item #8), derived from previous confirmatory factor analysis studies on BDI-II (48,49). Each item was rated on a four-point scale [0-3] with higher scores indicating greater symptom severity and treated as ordinal variables for logistic regression analysis. We additionally composed a global subscale score for both affective components, estimated as the total sum of item scores for each component (treated as a continuous variable) of the constituent items under each component. Higher sum scores indicated more severe anhedonia or increased pessimistic attributional style, respectively.

### Statistical methods

Associations between fMRI contrast weights and CRP were performed using one-sided linear regression within the group of all depression cases using the *lm* function in R (v3.6.0), and subsequently, *pt* function to estimate p-values for left-tailed (beta < 0; hypoactivation and negative scaling with CRP) and right-tailed (beta > 0; hyperactivation and positive scaling with CRP) respectively. Associations of fMRI activation with behavioral phenotypes were tested using ordinal logistic regression instrumentalized via the *polr* function in R (v3.6.0).

## Results

### Participant characteristics

After QC procedures, analyzable fMRI and CRP data were available for N=124 participants, comprising N=45 HC, N=48 low CRP depression cases and N=31 high CRP depression cases. Sociodemographic and clinical variables are summarized in **Table S2.** High CRP depression cases included proportionally more females and had higher BMI than low CRP depression cases. Depression cases were not excluded if currently prescribed antidepressants and/or pre-specified concomitant medication for minor diagnostic comorbidities.

### Functional activation in response to monetary outcomes

Monetary rewards (Win > No-win contrast for gain condition) were associated with significant activation within the vmPFC in healthy controls and all depression cases, with marginally higher peak activation in controls compared to cases (**Table 1; Figure 2**). However, when stratified categorically based on CRP concentration, significant activation of the vmPFC was only observed in low CRP depression cases and did not survive voxel-wise thresholding in high CRP depression cases, even under less conservative thresholding with FDR correction at 5%. Significant activation of the right ventral striatum (NAc) was only observed in cases of depression and not in healthy controls. This effect was again specific to low CRP depression cases when the group of depression cases was categorically stratified by the CRP < 3 mg/L criterion (**Table 1; Figure 2**). Between-group comparisons demonstrated no significant case-control differences in activation for either vmPFC or ventral striatum (NAc) (**Figure S3**).

**Table 1.**
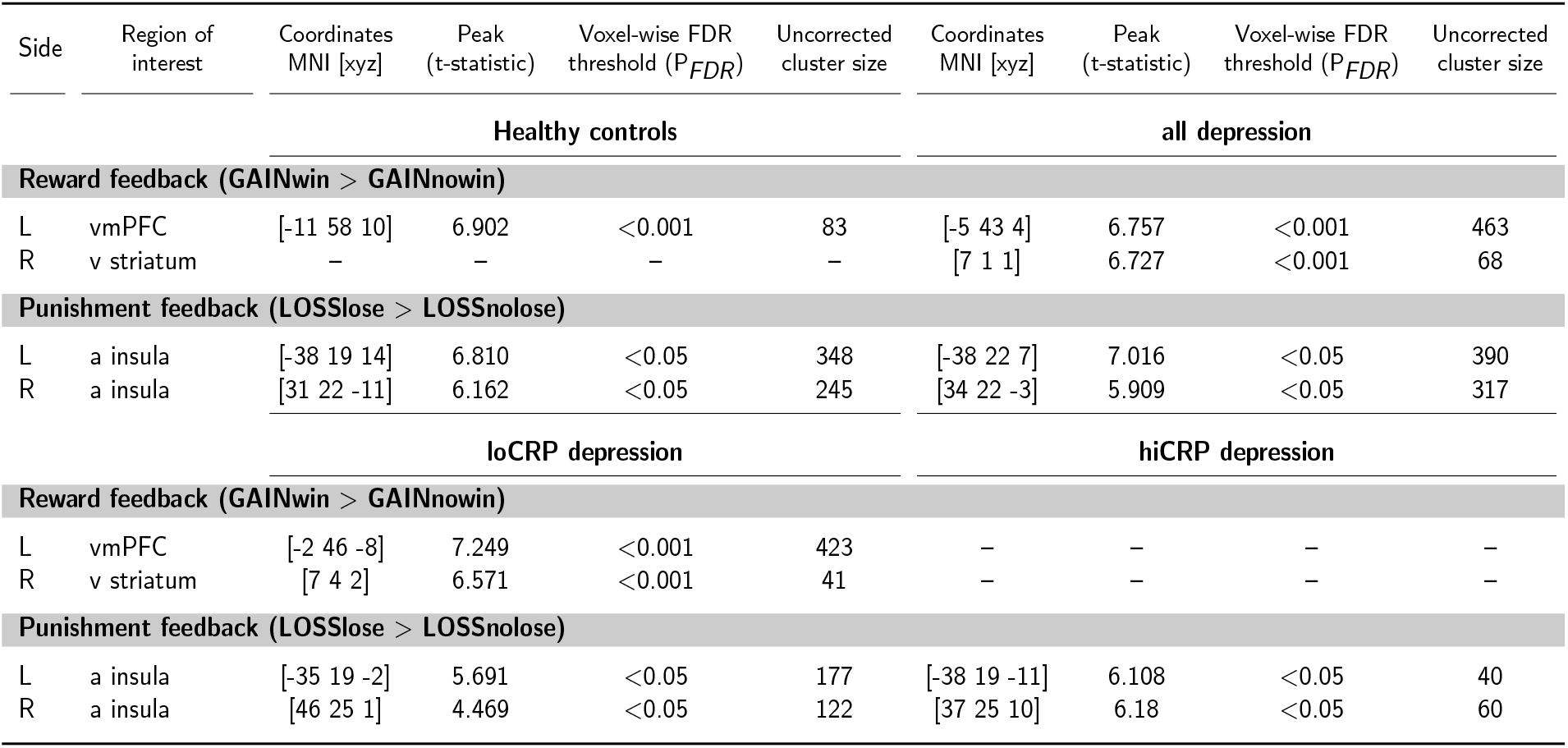
Local maxima of task-related fMRI activation for monetary reward (Gain condition) and punishment (Loss condition) outcome events. Significant voxel-wise functional activation by reward (monetary gain) and punishment (monetary loss) outcomes across groups. *loCRP depression; low CRP depression cases, hiCRP depression; high CRP depression cases*.

**Figure 2.**
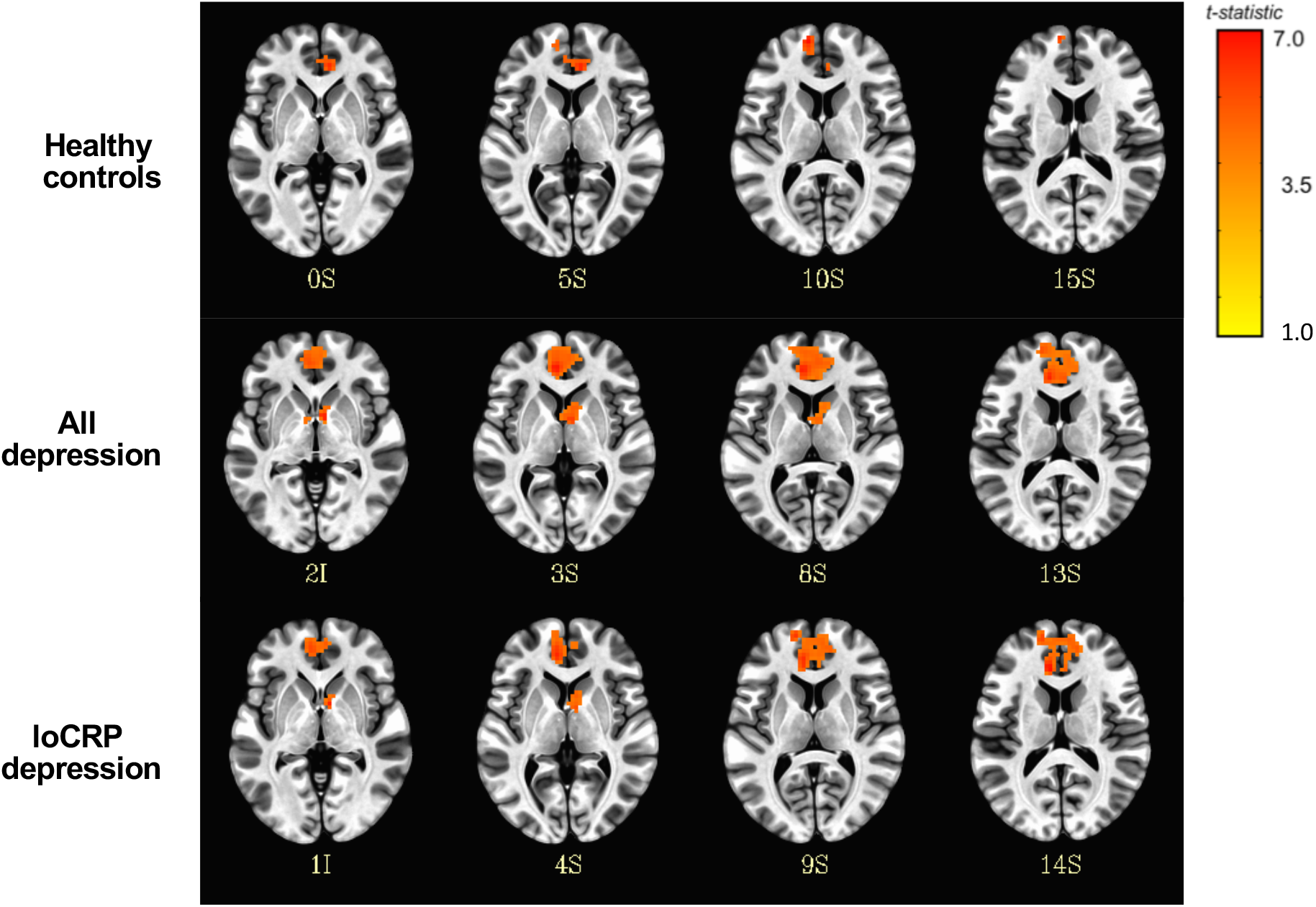
Functional activation to monetary reward outcome (Gain condition). Statistical parametric maps of significant vmPFC and striatal voxel-wise activation to monetary reward across groups. All activation maps are depicted in MNI space in neurological convention (left side of the brain on left side of the picture). The color bar depicts t-values of local activated voxels. *loCRP depression; low CRP depression cases*.

Monetary losses (Lose > No-lose contrast in the loss condition) significantly activated the left and right anterior insular regions across all four groups (**Table 1; Figure 3**). However, the experimental effect of punishment was generally weaker than the effect of reward, in that significant activation was only noted under less conservative voxel-wise thresholding after correction with FDR < 5%. Between depression subgroups, peak activation was higher in high CRP depression cases compared to low CRP depression cases. Between-group comparisons yielded no significant differences in punishment-related activation of bilateral anterior insula (**Figure S4**).

**Figure 3.**
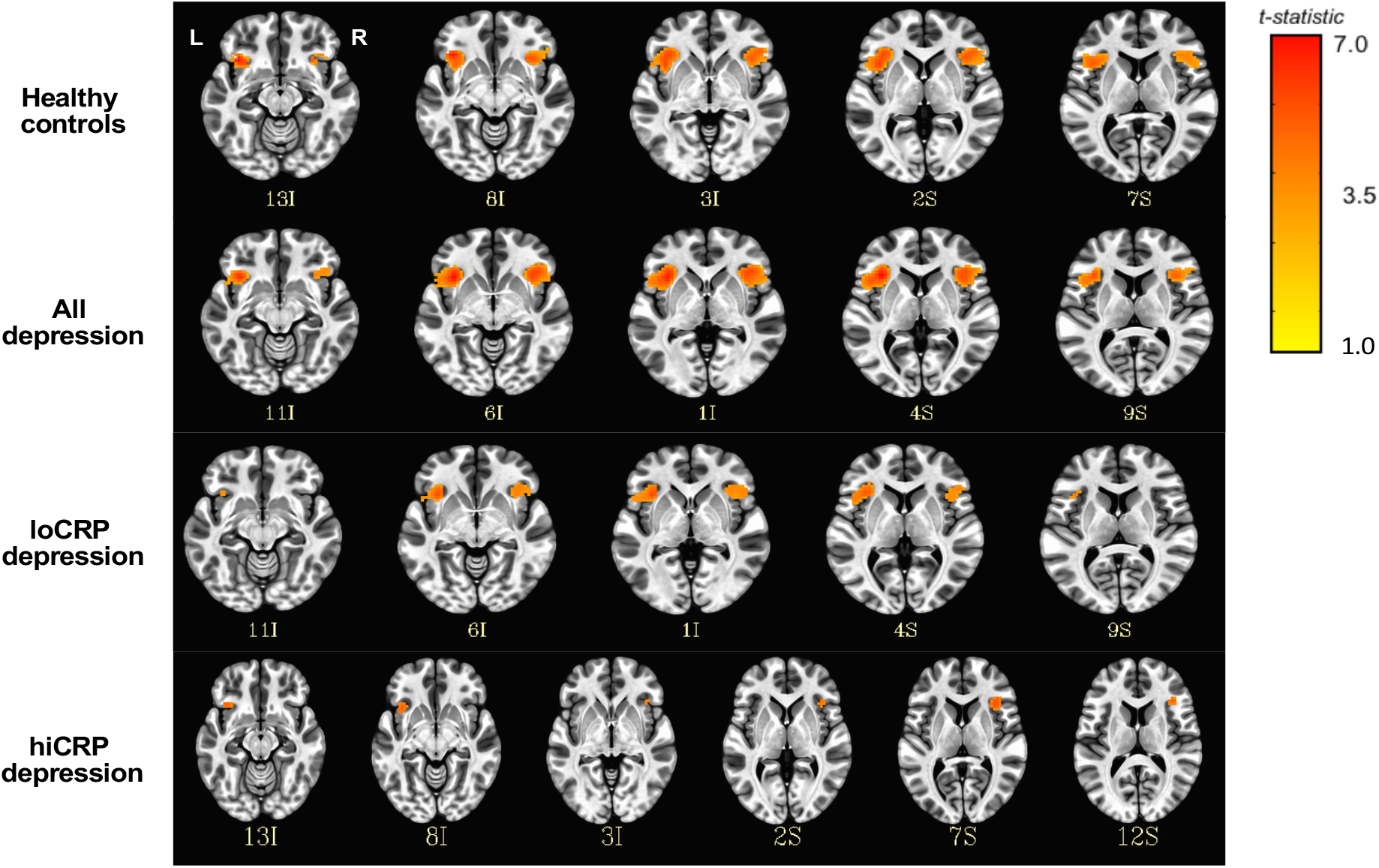
Functional activation to monetary punishment outcome (Loss condition). Statistical parametric maps of significant voxel-wise insular activation to monetary punishment across groups. All activation maps are depicted in MNI space in neurological convention. The color bar depicts t-values of local activated voxels. *loCRP depression; low CRP depression cases, hiCRP depression; high CRP depression cases*.

### Association between functional activation to monetary reward or punishment outcomes and peripheral inflammation

We extracted BOLD parameter estimates from atlas-derived masks of regions indicating activation to Win > No-win and Lose > No-lose contrasts previously examined i.e. vmPFC, ventral striatum (NAc) and anterior insula. For each region of interest (vmPFC, NAC, anterior insula), estimates of task-related activation from the left and right hemispheres were significantly correlated (vmPFC, *r* =0.78, *P* =2.2e-16; NAc, *r* =0.65, *P* =1.095e-10; anterior insula, *r* =0.62, *P* =2.101e-09), and since it is unclear whether reward and/or punishment functions are hemispherically lateralized (50), we *a priori* tested the selected ROIs laterally i.e. left and right separately.

As hypothesized, contrast weights extracted from the right vmPFC mask (*r* =-0.21; *Pone-sided* =0.035) and left NAc (*r* =-0.21; *Pone-sided* =0.039) negatively scaled with CRP. These results indicated lower levels of reward-related activation or hyposensitivity of these regions with increasing peripheral inflammation (**Figure 4 A-B**). However, contrary to our prior prediction, neither left (*r* = −0.04; *Pone-sided* = 0.37) nor right (*r* =0.1; *Pone-sided* =0.39) anterior insula showed positive scaling of punishment-related activation with increasing CRP (**Figure 4 C-D**).

**Figure 4.**
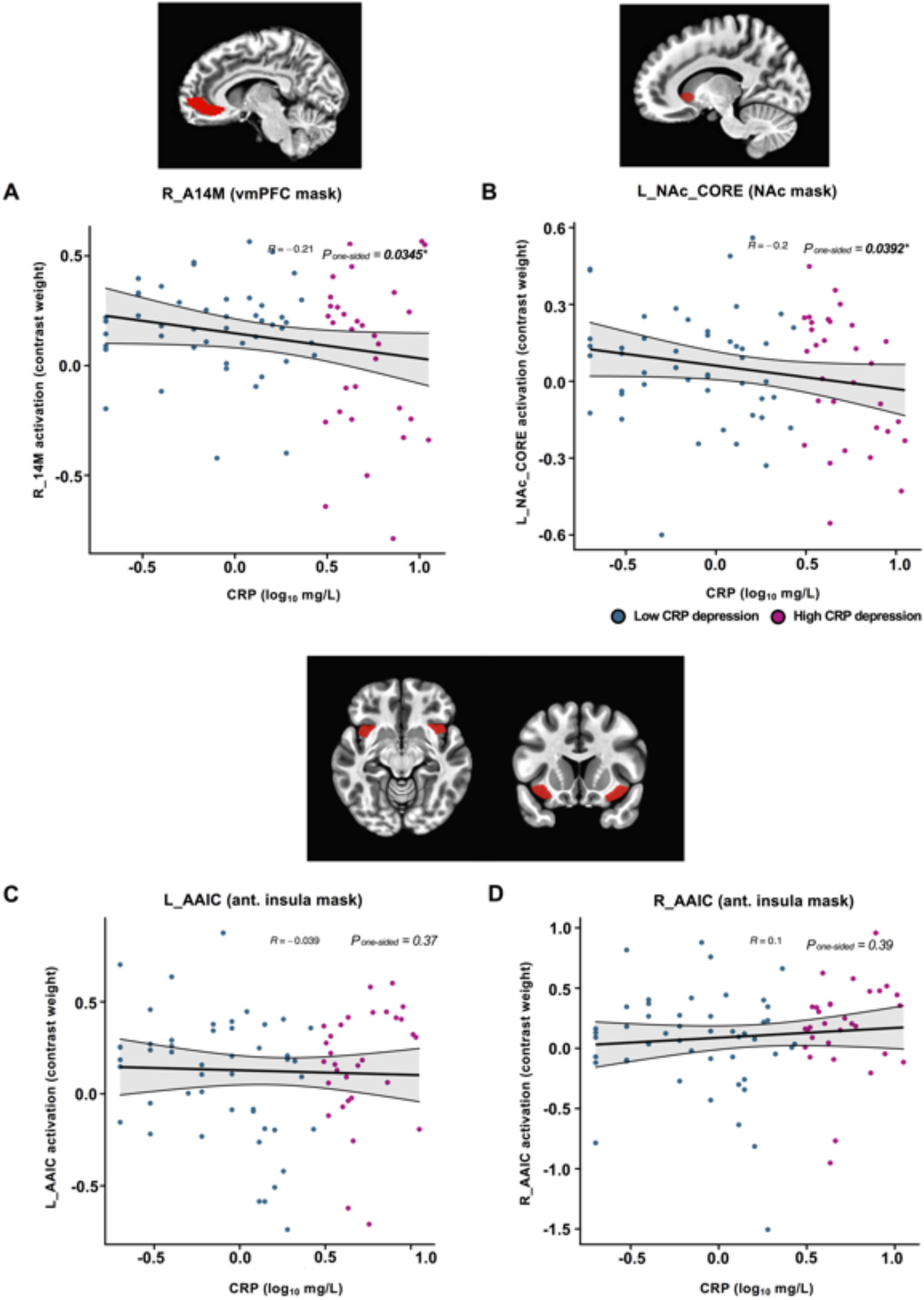
Association between blood CRP concentration and monetary reward-or punishment-related activation of selected brain regions. Reduced activation by reward outcomes in right ventromedial prefrontal cortex (**A**) and left nucleus accumbens (**B**) region of the ventral striatum was significantly associated with increasing CRP. There was no evidence for significant association between CRP and increased activation by punishing outcomes in the anterior insular regions (**C-D**).

### Association between functional activation and behavioral affective scores

Despite demonstrating negative scaling with inflammation, higher activation of the right vmPFC and left NAc were not significantly associated with more severe anhedonic attitudes (**Figure S5A; first and second columns**). Instead, higher activation of both regions showed non-significantly increased odds of more severe ‘loss of interest’ (vmPFC, 5% higher odds, OR =1.05, 95%CI 0.22 – 4.5; NAc, 81% higher odds, OR =1.81, 95%CI 0.279 – 11.77). Other anhedonic attitudes had non-significantly increased or decreased odds in association with heightened activation of the reward-related brain regions (**Figure S5A; first and second columns**). Corroborating this result, composite scores generated from the anhedonia-related items also did not demonstrate an association with vmPFC (*r* =-0.01; *P* =0.93) or NAc (*r* =0.01; *P* =0.91) activation (**Figure S5A; third column**).

Likewise, higher activation of the left and right anterior insula was also not significantly associated with higher odds of negative attitudes (**Figure S5B; first and second columns**). However, both regions showed a more consistent pattern of association, in that all negative behaviors showed at least nominal increase in odds ratio associated with activation of punishment-related brain regions, i.e. 1% higher odds of ‘punishment feelings’ to 63% of ‘pessimism’ in the left anterior insula, and 18% higher odds of ‘self-criticalness’ to 56% of ‘pessimism’ (**Figure S5B; first and second columns**). Composite scores were also not significantly associated with punishment-related activation of left (*r* =0.06; *P* =0.60) or right (*r* =0.09; *P* =0.44) anterior insula activation, although slopes for both regressions were positive suggesting a trend towards increased negative attitudes in association with hyperactivation of the insular regions by punishing trials (**Figure S5B, third column**).

## Discussion

Positive and negative affective abnormalities i.e. dichotomous alterations to positive and negative affect, are integral symptoms of depression. However, it is still not well elucidated if and how peripheral inflammation might modulate these features in depression. Our investigation yielded three key results.

First, we did not find any statistically significant differences in activation to either reward or punishment outcomes, between any two groups. Second, we tested a similar set of hypotheses on hyposensitivity to reward, and hypersensitivity to punishment, as a function of increasing peripheral inflammation, within depression cases only. As predicted, we showed that reward-related activation within the vmPFC and left NAc regions negatively scaled with CRP, suggesting hyposensitivity to reward in depression cases with increasing peripheral inflammation. However, contrary to our expectation, we did not observe any association between punishment-related activation within the anterior insula and CRP. Finally, brain functional activations to reward and punishment outcomes were not significantly associated with behavioral measures of anhedonic and negative attitudes, respectively. In sum, these observations partially corroborated the existing literature, showing support for desensitization of the neural reward system with increasing peripheral inflammation in depression but not reciprocal hypersensitization of the neural alarm system.

### Desensitization of the neural reward system but not hypersensitization of the neural alarm system in inflammation-linked depression

Down-regulation of ventral striatal (NAc core) responses to positively valenced outcome (monetary reward) as a function of systemic CRP corroborates several previous task fMRI reports on human inflammation-induced experimental models of depression (34,40,51). Although study setting, task paradigms and analytic design vary across studies, investigations have collectively shown that inflammation not only modulates ventral striatal responses to reward consumption or receipt e.g. via presentation of outcome or feedback on winnings (40), but also anticipation of reward (39,52).

However, a reciprocal up-regulation of anterior insula activation by negatively valenced outcome (monetary punishment) against CRP was not observed in our data, despite being more convincingly implicated in studies of exogenously-induced inflammation-linked depression, and demonstration of insula immune-encoding activity in a recent animal study (53). One study (34) had demonstrated hypersensitivity of the anterior insula to similar punishment signals, i.e., monetary loss in the context of inflammation and depression.

Another (54) also showed a positive scaling between bilateral anterior insula activation to social rejection (Cyberball task) and TNFαRII following exposure to laboratory social stressors. Nonetheless, this was an experimental sample of healthy immunologically unchallenged individuals with salivary inflammatory markers and not peripheral blood-derived. In reconciling this surprising finding, it is worth recognizing that punishing stimuli come in many forms. Monetary loss, grief, social threat, social stress, pain, social defeat and negative feedback from an “evaluator” are all punishing. But whether these are encoded similarly within the brain, is still largely unclear (55,56).

### Inflammation enhances generalized interoceptive dysfunction in depression

An alternative view is that the combination of these observations i.e. attenuated response to reward and heightened response to punishment is consistent with the emotional context insensitivity hypothesis (ECI) (57,58). Under this model, depression is characterized by blunted response or reactivity to all emotional stimuli regardless of valency. In other words, it holds that those with depression exhibit neutral reactivity to experiences of both pleasantness and unpleasantness, nothing is of extreme positive or negative valence. This contrasts with the perhaps more ubiquitously held framework on depression, that is, the dichotomous coupling of reward system hyposensitization and threat or alarm system hypersensitization. Nonetheless, it agrees with observations of the current study, where critically, (i) punishment associated neural signal was generally diminished compared to reward-related neural activation in depression cases (**Table 1**), and (ii) no scaling was observed between anterior insular activation and inflammation indexed by CRP (**Figure 4**). Also supporting the ECI hypothesis, an early case-control meta-analysis on emotional reactivity in depression (59) reported a generalized reduction in emotional activity to both positively and negatively valenced stimuli in MDD (although the effect size was slightly larger for positive stimuli than for negative stimuli).

Interestingly, the ECI view overlaps heavily with an analogous contemporary framework known as interoception i.e. perception of internal bodily sensations. Interoception transcends both the biomedical and psychological conceptualizations of depression. Key features of interoceptive abnormalities include alexithymia or apathy (60,61), which is closely analogous to emotional insensitivity in the ECI literature. Our previous finding on a larger subset of the present sample (41), indeed suggested an interoceptive nervous system dysfunction (reduced connectivity within the network) with increasing peripheral inflammation. These results converged with prior reports of insular functional MRI activation changes in inflammation-linked depression (62).

### Strengths and limitations

A key strength of our study was principally in the case-control study design for whole-brain voxel-wise study of affective reorientation in inflammation-linked depression. Although the extant body of work has thematically explored this question, methodologically, prior results were mainly from human experimental models of inflammation-linked depression where the inflammatory condition was experimentally controlled or “manipulated”, e.g. by LPS endotoxemia immune challenge, typhoid vaccination or IFNα immunotherapy (17,35,63); as opposed to an “observational” study where uncontrolled peripheral inflammation is assessed, such as the present study. This is important to distinguish in the context of inflammation-linked depression as the nature of peripheral inflammation in this subgroup of depression cases is typically chronic and low-grade. Therefore, it is conceivable that extent and construct of affective abnormalities, especially reorientation in neural response, may be different i.e. perhaps modest in effect size, compared to those reported via experimental challenge studies.

An important limitation of our current study is the lack of high CRP (> 3mg/L) control group. In future, it will be important also to analyze differences between depression cases and non-depression controls, with CRP levels allowed to exceed the 3 mg/L cut-off in both groups, to determine if functional activation to reward and punishment is similarly modulated in a non-depression population with chronic low-grade peripheral inflammation. It is also worth noting that the small effect size and surprising null observation for an association insula activation to punishment and CRP, could equally be confounded by poor task comprehension and performance by participants drawn from a clinical population. As part of task performance validation and quality control, we examined (i) overall performance or task accuracy (**Figure S6**), (ii) trial-by-trial learning for each condition (**Figure S7**), (iii) learning rate (α) and exploration/exploitation trade-off (β) model parameters derived from learning algorithm (**Figure S8-9**), and (iv) functional activation to cue or stimuli pair presentation i.e. Gain > Look; Loss > Look (**Figure S10-12**). Although effort was made to ensure participants understood the task instructions, findings from additional QC indicated that some participants likely did not fully comprehend the task.

## Conclusions

While reorientation to rewards and punishments have been heavily evidenced in depression, and to a lesser extent in human experimental models of inflammation-linked depression, to our knowledge, this study provides the first evidence for an association between increasing inflammation and hyposensitivity to reward within the vmPFC and NAc in a depression sample with low-grade peripheral inflammation; and, unexpectedly, a lack of association to punishment within the anterior insula. The findings partially replicate the prevailing understanding on brain functional impairments in inflammation-linked depression, i.e., reward system attenuation.

## Data Availability

All data produced in the present study will be made available upon reasonable request to the authors following peer-reviewed publication.

## Acknowledgements

The authors thank all the participants in the study and members of the NIMA research team – in particular, project coordinators Linda Pointon, Junaid Bhatti, Ciara O’Donnell (see **Supplementary Appendix** for a complete list of NIMA Consortium members), Dr Lorinda Turner (Cambridge Biomedical Research Centre) for processing and assaying blood samples, medical imaging staff and laboratory staff.

