## Supplemental Data (SD) for "Reward-related activation of fronto-striatal regions scaled negatively with C-reactive protein but showed no association with anhedonia in depression"

[§]full list of authors available in the Supplementary Appendix.

*these authors contributed equally to this work and share last authorship.

**Correspondence**

Miss Athina Aruldass

Level 5 (Room 515), Department of Psychiatry,

Clifford Allbutt Building, Cambridge Biomedical Campus,

CAMBRIDGE, CB2 0AH, UK.

### Contents

#### *Supplementary Methods*


| | | |
|---|---|---|
| **Section 1** | **Participants** | **3** |
| | **Figure S1** Schematic of quality control and analytic pipeline | **3** |
| **Section 2** | **Immunophenotyping** | **4** |
| **Section 3** | **Computational modelling of observed behavior** | **5** |
| | **Figure S2** Algorithms used to model observed behavior | **6** |
| **Section 4** | **fMRI acquisition and preprocessing** | **7** |
| **Section 5** | **fMRI confirmatory analyses** | **7** |
| | **Table S1** Regions-of-interest (ROI) masks derived from atlases for confirmatory analyses | **7** |


#### *Supplementary Results*


| | | |
|---|---|---|
| **Section 6** | **Sample characteristics** | **8** |
| | **Table S2** Quality-controlled analyzable cohort of task-related fMRI data (N=124) | **8** |
| **Section 7** | **Case-control differences in functional activation** | **9** |
| | **Figure S3** Between-group comparison for difference in activation to rewarding outcomes | **9** |
| | **Figure S4** Between-group comparison for difference in activation to punishing outcomes | **10** |
| **Section 8** | **Association between functional activation and affective scores in depression** | **11** |
| | **Figure S5** Association between neural activation and behavioral attitudes in depression cases | **11** |
| **Section 9** | **Quality control outcomes** | **12** |
| | **Figure S6** Accuracy of response per trial type and overall performance | **13** |
| | **Figure S7 Trial-by-trial learning curves for each condition** | **13** |
| | **Figure S8** Computational model parameters α (learning rate) and β (exploitation- exploration index) | **15** |
| | **Figure S9** Motor performance (button pressing) pattern | **16** |
| | **Figure S10** Neural activation to button pressing (choice) event | **17** |
| | **Figure S11** Functional activation to presentation of Gain stimuli (GAIN > LOOK contrast) | **17** |
| | **Figure S12** Functional activation to presentation of Loss stimuli (LOSS > LOOK contrast) | **18** |

**References** **19**

### *Supplementary Methods*

#### Section 1 Participants and overall analytic and quality control pipeline

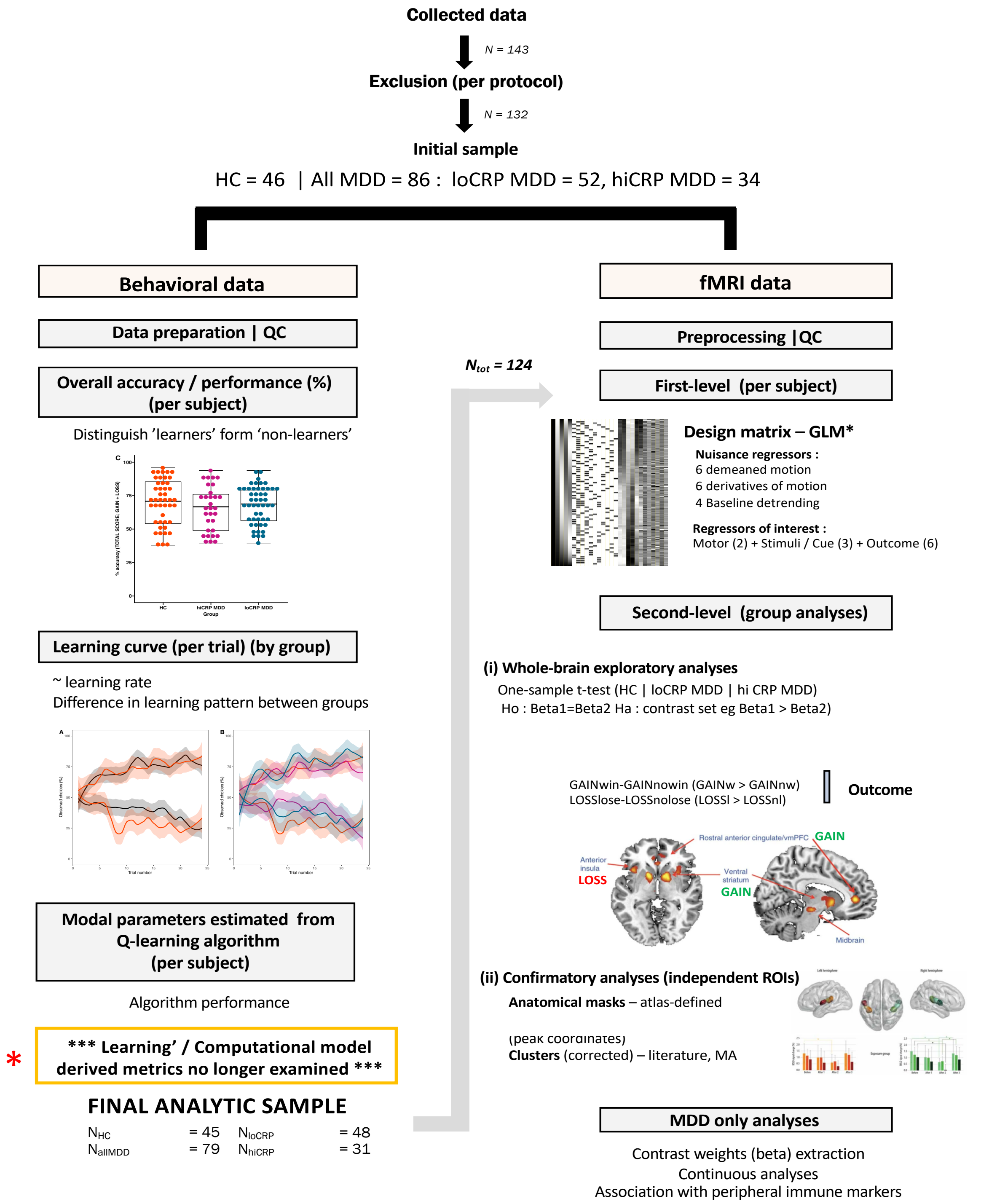


**Figure S1 Schematic of quality control and analytic pipeline.** Quality control outcomes of behavioral and imaging data are detailed in *Section 9*.

### Section 2 Immunophenotyping

*Serum and plasma collection*

We collected up to 90mL of venous blood by antecubital venepuncture from participants who had fasted overnight from midnight to between 08:00 and 10:30am on the day of assessment. Participants had refrained from exercise for 72 hours to blood sampling. Blood (8.5ml) were collected into BD Vacutainer® Serum Separator Tubes (SST™ II Advance) Silica tubes and Plasma Preparation Tubes (PPT™) and incubated for 30 minutes at room temperature to allow blood to coagulate. Tubes were centrifuged at 1600 Relative Centrifugal Force (rcf) for 15 minutes and serum or plasma collected in aliquots. All serum samples were stored at -70°C until analysis, with no freeze-thaw cycles.

*high-sensitivity CRP assay*

Collected blood samples were allowed to coagulate for 30-60 minutes then centrifuged at 1600rcf for 15 mins. 1ml of the resultant sample was then transferred to a white-topped serum tube using a pipette and transported at room temperature to central laboratory ($Q^2$ solutions). Samples were exposed to anti- CRP-antibodies on latex particles, and the increase in light absorption due to complex formation was used to quantify CRP levels, using Turbidimetry on Beckman Coulter AU analyzers. Both initial stratification hsCRP assay and MRI appointment hsCRP assay were performed at a single central laboratory ($Q^2$ Solutions, The Alba Campus, Livingston EH54 7EG, UK) from 0.5mL of plasma.

### Section 3 Computational modelling of observed behavior

To model learning behavior computationally, we fitted trial-by-trial task performance to a standard reinforcement learning algorithm (**Figure S2**). Choice values were updated with a simple Q-learning algorithm.

Expected values ($Q_A$ and $Q_B$) were initialized at 0 and the value of the stimulus chosen at each trial (e.g. option A) was updated according to the rule:

$Q_{A(t + 1)} = Q_{A(t)} + \alpha \cdot \delta_{(t)}$ , with outcome prediction error δ at trial *t* defined as the difference between the actual outcome (R) and expected ($Q_A$) outcome ; $\delta_{(t)} = R_{(t)} – Q_{A(t)}$

The probability of choice selection was then estimated with a softmax decision rule Given the expected values ($Q_A$) and ($Q_B$) at a trial *t*, what is the probability of a subject selecting option A i.e. P(A) at trial *t*. This was calculated using the function:

$P(A)_{(t)} = \exp(Q_{A(t)} / \beta ) / \exp[Q_{A(t)} / \beta ] + \exp[Q_{B(t)} / \beta ]$

The free parameters α (learning rate) and β (temperature or exploration:exploitation or degree of randomness in selection making) were adjusted to maximize the likelihood of each participant’s observed choices under the model. *R was defined as the* actual outcome, with magnitudes encoded as +£1, £0 for ‘nothing’, or -£1).

Altogether, the algorithm iteratively sampled α and β parameters under a search grid ranging from 0 to 1, to determine the optimal pair that best estimated the actual observed choices of each individual. The log likelihood was determined for each combination of α and β parameters (between 0 and 1) for each set of trials. The optimal pair of α and β parameters was determined to be the pair corresponding to the maximum log likelihood (highest cumulative log likelihood i.e. sum across set of trials) for each individual. The algorithm was coded using MATLAB.

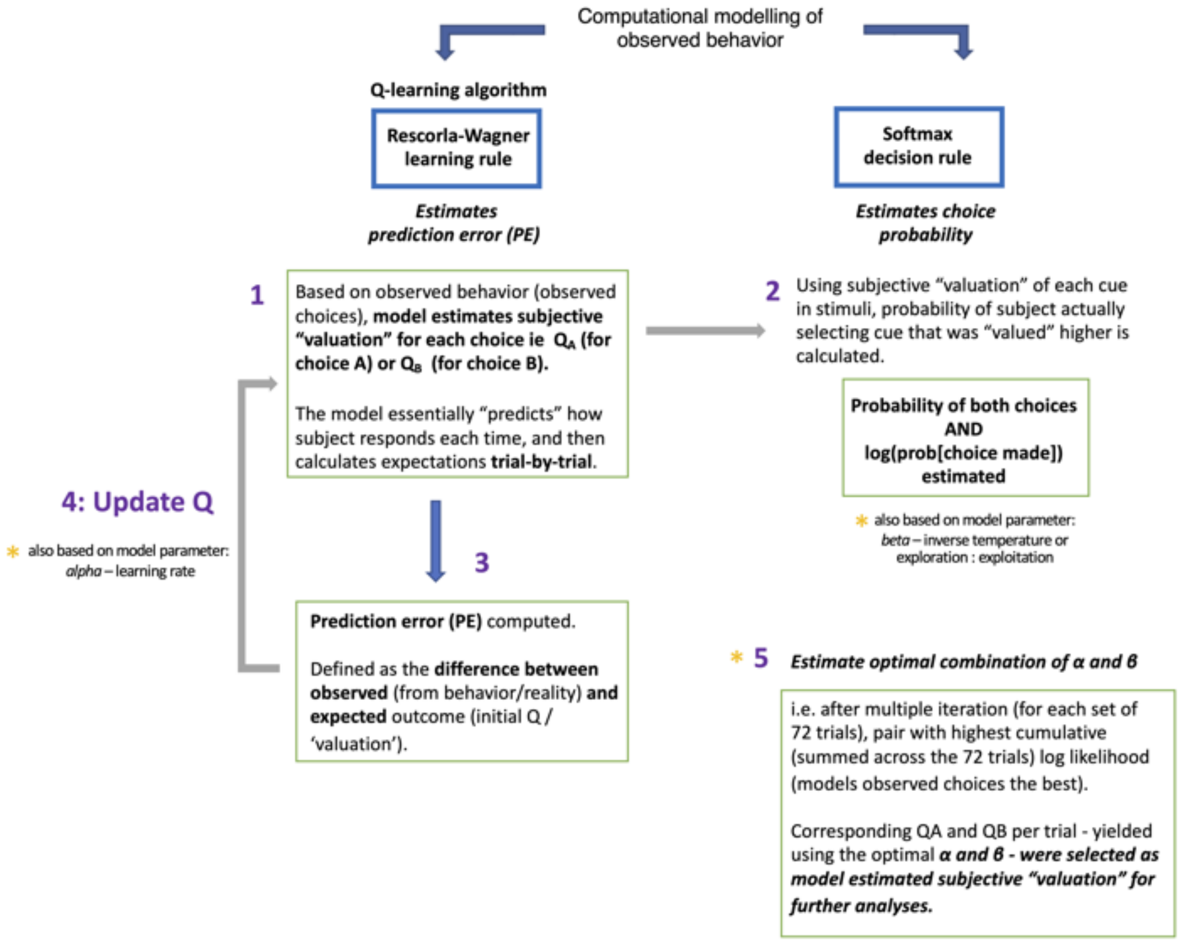


**Figure S2 Algorithms implemented to estimate model-based behavioral choices during probabilistic stimulus task.**

#### Section 4 fMRI acquisition and preprocessing

*fMRI data acquisition*

Functional scans were acquired using an echo planar imaging (EPI) sequence with the following parameters: relaxation time (TR) = 2.52s; echo times (TE) = 30 / 43ms (site dependent); acquisition time = 10.1 minutes = 241 time points. Data were collected as 34 slices at -30 degrees to the AC-PC line with field of view (FoV) = 192 mm and matrix size = 64 × 64, for voxel resolution of 3.75 x 3.75 x 3.99mm. Images were projected onto a screen at the foot of the participant. Participants viewed the stimuli via a mirror placed above the top of the head coil reflecting images from the screen.

*fMRI data preprocessing*

Preprocessing of task-based fMRI data was performed with AFNI (version 21.0) using the *afni_proc.py* script (4). This pipeline comprises the following standard fMRI preprocessing steps: slice time correction, despiking, volume registration, alignment to the T1 anatomical image, masking, spatial smoothing and scaling. The first 5 images were discarded as dummy scans (to allow for magnetization to reach steady state) and functional images were corrected for slice-time acquisition and realigned to adjust for motion-related artifacts. All images were spatially smoothed with a full-width at half-maximum (FWHM) 8.5mm Gaussian kernel. The corrected images were spatially registered to the T1 anatomical data, normalized to the Montreal Neurological Institute (MNI) template (MNI152), and resampled as 2.68mm$^3$ voxels. For movement correction, first-level timeseries regression was performed after computing (i) demeaned motion parameters, (ii) derivatives of motion parameters, and (iii) volumes to censor. Images with large head motion (default threshold set at 0.3mm between successive time points (framewise displacement), based on the motion parameters), as well as censoring of outlier time points, where at least 5% of the brain voxels are computed as outliers.

#### Section 5 Confirmatory analyses ROIs

| Source | Feedback condition | Anatomical region | ROI name as identified by atlas) | Side | Centroid MNI coordinates [xyz] | Mask size (voxels) |
|---|---|---|---|---|---|---|
| **Atlas-derived mask** | | | | | | |
| **Tian et al.(2020) subcortical atlas** | Reward | ventral striatum | NAc-core | L | [-13 19 -3] | 848 |
| | Reward | ventral striatum | NAc-core | R | [15 19 -3] | 848 |
| **Fan et al.(2016) Brainnetome cortical & subcortical atlas** | Reward | vm PFC | A14m | L | [-7 54 -7] | 4116 |
| | Reward | vm PFC | A14m | R | [6 47 -7] | 5187 |
| **Glasser et al.(2016) cortical atlas** | Punishment | anterior insula | AAIC | L | [-35 14 -12] | 1859 |
| | Punishment | anterior insula | AAIC | R | [33 14 -14] | 1691 |

**Table S1 Regions-of-interest (ROI) masks derived from atlases for confirmatory analyses of depression- and inflammation-related effects on monetary reward or punishment.**

### *Supplementary Results*

### Section 6 Participant characteristics

| | HC (N=45) | | all depression (N=79) | | Case-Control difference | loCRP depression (N=48) | | hiCRP depression (N=31) | | loCRP–hiCRP difference |
|---|---|---|---|---|---|---|---|---|---|---|
| | Median | IQR | Median | IQR | *p-value* | Median | IQR | Median | IQR | *p-value* |
| **Sociodemographic / Clinical** | | | | | | | | | | |
| Age (years) | 35.2 | 11.6 | 38.9 | 12.4 | 0.18 | 39.0 | 11.5 | 36.3 | 11.6 | 0.73 |
| BMI ($kg/m^2$)[a] | 23.5 | 5.3 | 26.7 | 5.9 | < **0.001***** | 25.5 | 4.5 | 27.8 | 7.5 | < **0.01**** |
| Sex, Male (n, %) | 19 | 42.2 | 27 | 34.8 | 0.48 | 21 | 43.8 | 6 | 19.4 | < **0.05*** |
| Tobacco, current smokers (n, %) | 0 | 0.0 | 15 | 18.9 | < **0.01**** | 11 | 22.9 | 4 | 12.9 | 0.42 |
| Alcohol, current users (n, %) | 23 | 51.1 | 44 | 55.7 | 0.76 | 28 | 35.4 | 16 | 51.6 | 0.72 |
| Ethnicity, White (n, %) | 30 | 66.7 | 64 | 81.0 | 0.47 | 41 | 85.4 | 23 | 74.2 | 0.40 |
| **Behavioral Instruments** | | | | | | | | | | |
| HAM–D (17–items)[b] | 0 | 1 | 17 | 7 | < **0.001***** | 17 | 8 | 18 | 5.5 | 0.93 |
| BDI–II[c] | 1 | 3 | 25 | 10.5 | < **0.001***** | 25 | 10.3 | 26 | 9.5 | 0.55 |
| SHAPS[d] | 0 | 0 | 5 | 5 | < **0.001***** | 6 | 3.5 | 4 | 6 | 0.26 |
| CFS[e] | 11 | 1 | 20 | 7 | < **0.001***** | 20 | 7 | 20 | 6.5 | 0.73 |
| STAI – S (items 1 - 20)[f] | 25 | 9 | 51 | 13.5 | < **0.001***** | 52.5 | 14.5 | 49 | 10.5 | 0.43 |
| STAI – T (items 21 - 40)[f] | 28 | 9 | 61 | 11.5 | < **0.001***** | 61 | 11.25 | 61 | 12 | 0.98 |
| CTQ[g] | 37 | 5 | 51 | 24 | < **0.001***** | 53 | 20.5 | 46 | 15.5 | < **0.05*** |
| PSS[h] | 12 | 8 | 26 | 6 | < **0.001***** | 26.5 | 5.3 | 26 | 6 | 0.89 |
| LEQ (score)[i] | 0 | 1 | 1 | 2 | < **0.001***** | 1 | 2 | 1 | 1 | 0.52 |
| LEQ (rating)[i] | 1.5 | 1 | 3 | 4 | < **0.001***** | 3 | 4 | 3 | 3.5 | 0.56 |
| **Inflammatory Biomarkers** | | | | | | | | | | |
| CRP (mg/ L)[j] | 0.6 | 1.1 | 1.8 | 3.5 | < **0.001***** | 0.9 | 1.0 | 4.5 | 3.6 | < **0.001***** |
| CRP ($log_{10}$ mg/L)[j†] | -0.2 | 0.4 | 0.2 | 0.5 | < **0.001***** | -0.1 | 0.3 | 0.7 | 0.2 | < **0.001***** |

**Table S2 Sociodemographic characteristics, questionnaire outcomes and serological features in quality-controlled analyzable cohort of task-related fMRI data (N=124).**
[a]Group differences were estimated using the Mann–Whitney U test or chi–squared test. body mass index; 1 HC missing data omitted in case-control statistical comparison; [b]Hamilton Rating Scale for Depression; [c]Beck's Depression Inventory (version II); [d]Snaith-Hamilton Pleasure Scale; [e]Chalder Fatigue Scale; [f] State-Trait Anxiety Inventory; [g]Childhood Trauma Questionnaire; [h]Perceived Stress Scale; [i]Life Events Questionnaire; [j] high-sensitivity C-reactive protein. IQR; interquartile range (Q3–Q1); †statistical comparison performed using unpaired t-test; **p* < .05; ***p* < .01; ****p* < .001 .

### Section 7 Between-group differences in functional activation to reward and punishment outcomes

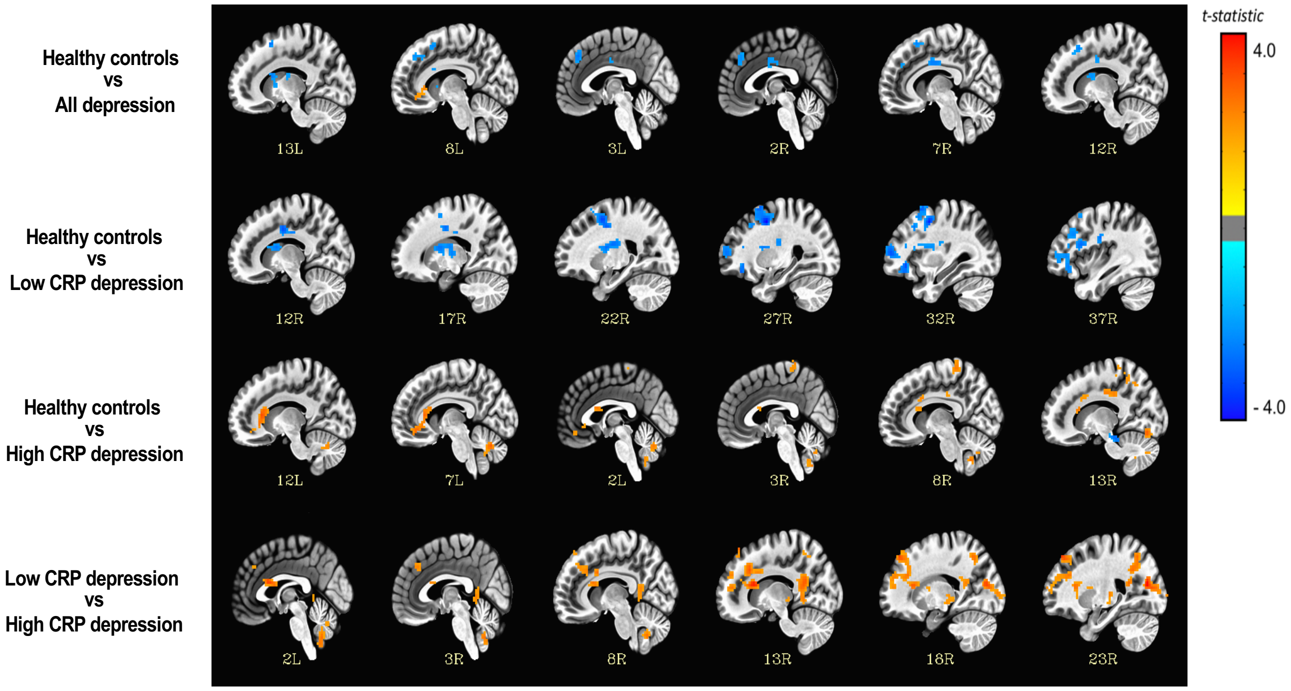


**Figure S3 Between-group comparison for difference in activation to rewarding outcomes.** No significant difference between any two groups for activation to monetary gains. All activation maps are depicted in MNI space in neurological convention (left side of the brain on left side of the picture). The color bar depicts t-values of local activated voxels.

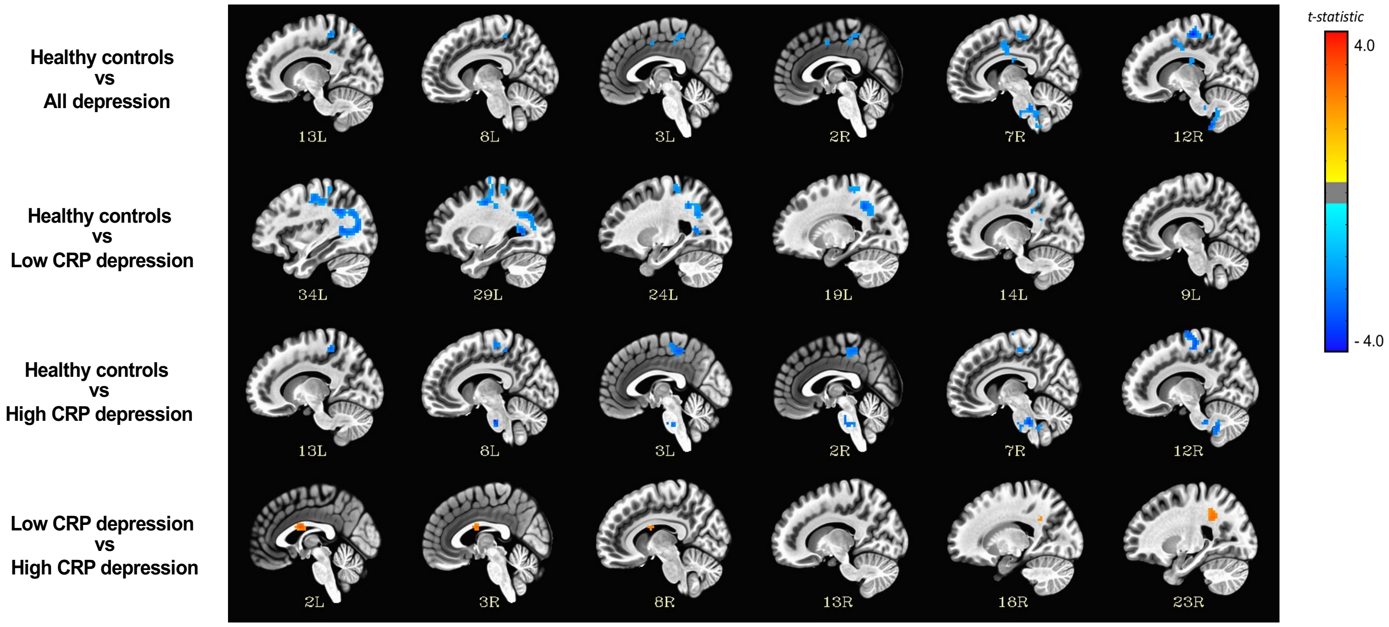


**Figure S4 No significant difference between any two groups for activation to punishing outcomes.** No significant difference between any two groups for activation to monetary losses. All activation maps are depicted in MNI space in neurological convention (left side of the brain on left side of the picture). The color bar depicts t-values of local activated voxels.

### Section 8 Association between functional activation and behavioral affective scores

**A**

| Dependent variable | Odds ratio | *P*-value | CI [lower, upper] |
|---|---|---|---|
| Loss of pleasure | 1.481 | 0.647 | [0.276,7.959] |
| Loss of interest in sex | 1.04 | 0.958 | [0.241,4.497] |
| Loss of interest | 1.053 | 0.948 | [0.224,4.96] |
| Loss of energy | 0.316 | 0.148 | [0.293,2.363] |

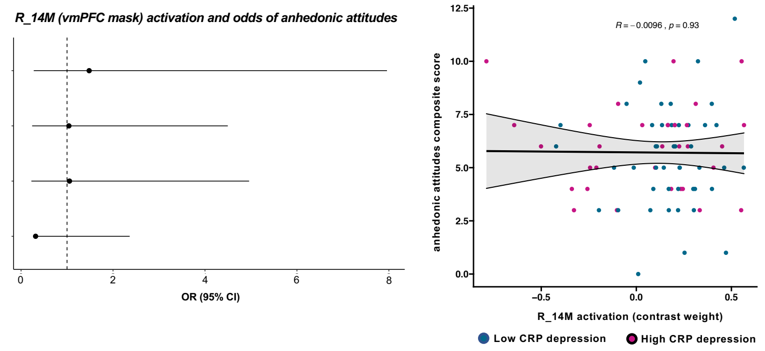


| Dependent variable | Odds ratio | *P*-value | CI [lower, upper] |
|---|---|---|---|
| Loss of pleasure | 0.8 | 0.824 | [0.113,5.682] |
| Loss of interest in sex | 0.605 | 0.563 | [0.11,3.325] |
| Loss of interest | 1.814 | 0.533 | [0.279,11.774] |
| Loss of energy | 1.159 | 0.881 | [0.168,7.977] |

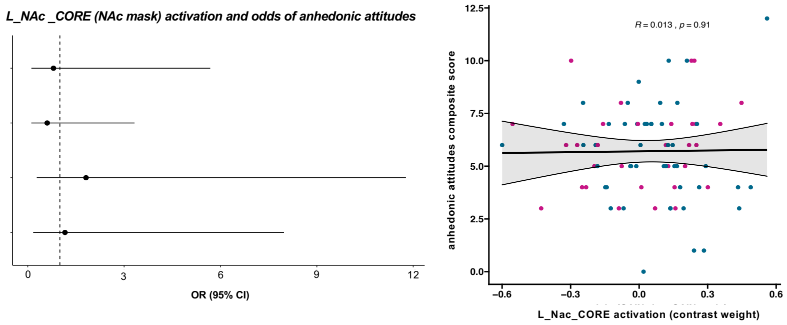


**B**

| Dependent variable | Odds ratio | *P*-value | CI [lower, upper] |
|---|---|---|---|
| Self dislike | 1.017 | 0.978 | [0.3,3.454] |
| Self criticalness | 1.374 | 0.61 | [0.405,4.658] |
| Punishment feelings | 1.01 | 0.989 | [0.254,4.016] |
| Pessimism | 1.628 | 0.458 | [0.45,5.891] |

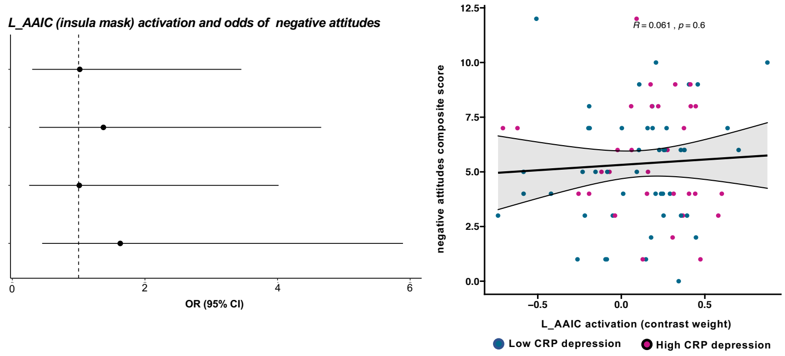


| Dependent variable | Odds ratio | *P*-value | CI [lower, upper] |
|---|---|---|---|
| Self dislike | 1.3 | 0.599 | [0.489,3.457] |
| Self criticalness | 1.179 | 0.743 | [0.441,3.147] |
| Punishment feelings | 1.225 | 0.714 | [0.414,3.623] |
| Pessimism | 1.56 | 0.392 | [0.564,4.317] |

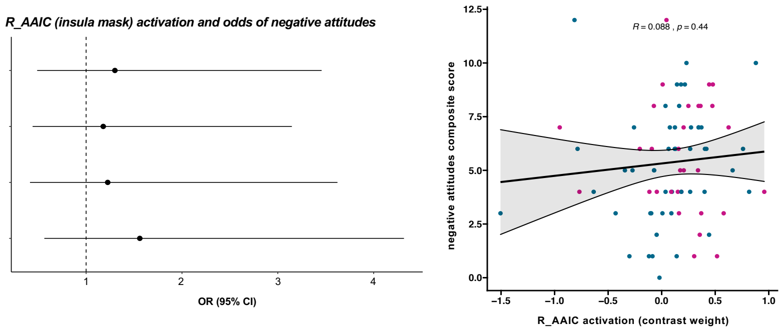


**Figure S5 Association between (A) reward-related neural activation and anhedonic attitudes and (B) punishment-related neural activation and negative attitudes.**

### Section 9 Quality control outcomes

*Computational model and fMRI task data quality control*

To ascertain the validity of the computational model and fMRI task data, that is, to examine whether the implemented task was reliably inducing the desired experimental effects, I performed QC to examine learning behaviour of participants through behavioral parameters estimated from the Q-learning algorithm and the fMRI data. This assessment of task validity is principally to ensure participants understood the probabilistic reinforcement learning task and were learning stimulus pairs which would consequently activate corresponding brain regions in response to anticipation of monetary reward (Gain condition) and punishment (Loss condition). Specifically, we examined (i) overall performance or task accuracy, (ii) trial-by-trial learning for each condition, (iii) learning rate (α) and exploration:exploitation (β) model parameters derived from learning algorithm, and finally (iv) functional activation to cue or stimuli pair presentation i.e. Gain > Look ; Loss > Look.

*(I) Behavioral task accuracy and performance*

Across the three groups and different trial types, a number of participants performed below chance, that is, scored < 60 % (observed correct choice) for each condition and cumulatively (Gain + Loss correct choices). However, more outlier participants were observed for the Gain condition than the Loss condition. No significant between-group differences were noted for either trial type (**Figure S6**) or total score (Gain + Loss correct choices), apart from marginally significant differences between healthy controls and all depression cases ($P_{\text{uncorrected}}$ = 0.041, Cohen's $d$ = 0.38), as well as between healthy controls and high CRP depression cases ($P_{\text{uncorrected}}$ = 0.046, Cohen's $d$ = 0.46), for the Loss condition. Trial-by-trial learning curves corroborated these findings and also provided the first indicator of task comprehension issues in participants especially upon examination of the last 50% of trials (**Figure S7**). For both conditions, trial-by-trial accuracy patterns were noticeably still erratic as opposed to stably diverging with greater time on task, i.e. consistently decreasing % of observed *incorrect* choices for the Loss condition and consistently increasing % of observed correct choices for the Gain condition. Similarly, no significant differences in learning curves were noted between groups for each trial type (**Figure S7**), apart from between healthy controls and all depression cases ($P_{\text{uncorrected}}$ = 0.029, Cohen's $d$ = 0.65), as well as between healthy controls and high CRP depression cases ($P_{\text{uncorrected}}$ = 0.011, Cohen's $d$ = 0.76) for the Loss condition.

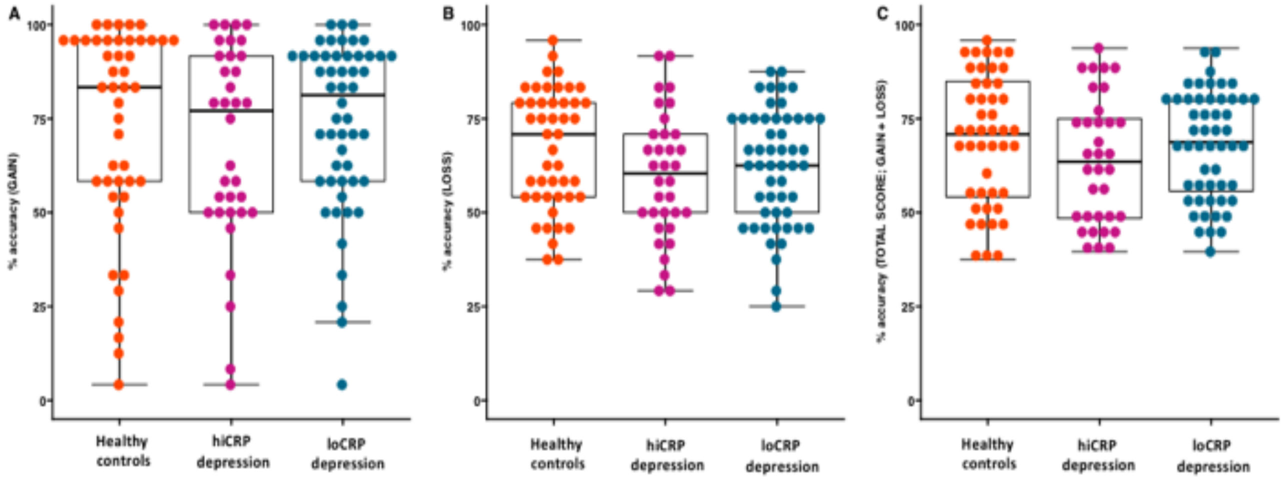


**Figure S6 Accuracy of response per trial type, including total score for overall performance**. A number of participants were noted to perform below chance, < 60% for each condition.

*(II) Trial-by-trial learning performance*

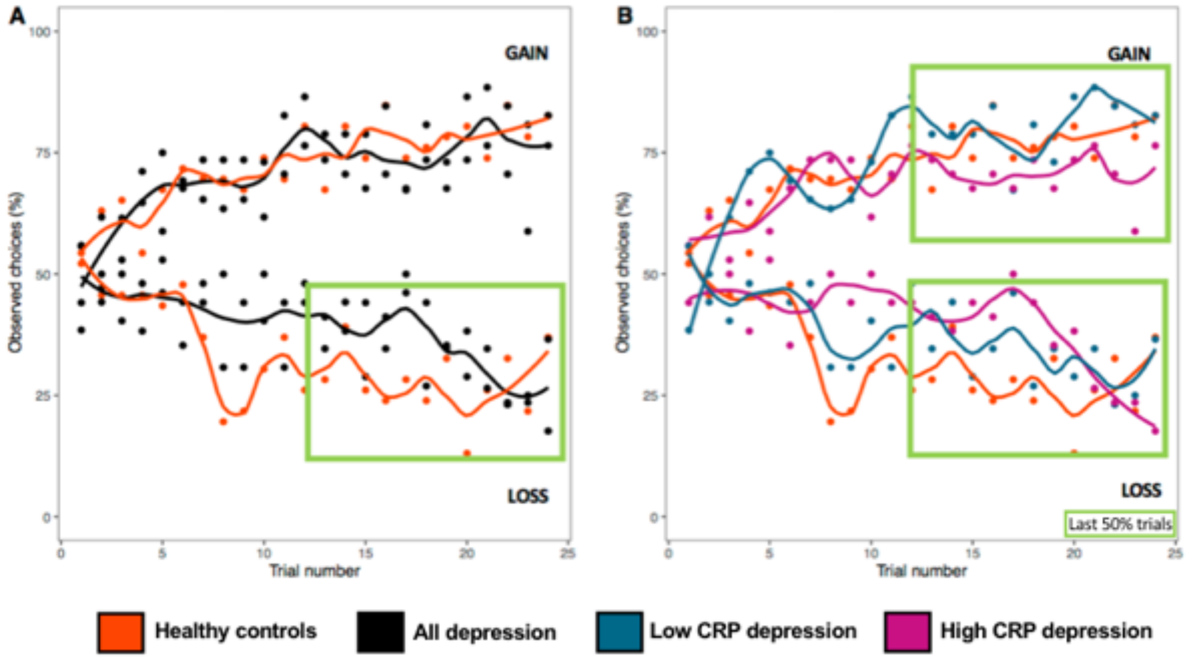


**Figure S7 Trial-by-trial learning curves for each condition in (A) all cases vs controls and (B) 3-groups (controls and CRP sub-stratified depression cases).** Observed choices – for Gain condition, % of 'correct' choice per trial, and for Loss condition, % or 'incorrect' choice per trial. For Loss condition, difference in learning is more evident at last 50% of trials (green box) between groups, where less incorrect answer is observed with HC least < low CRP cases < high CRP cases. For Gain condition, minimal difference in learning between groups at last 50% of trials, especially between low CRP and high CRP depression subgroups. However, performance was notably erratic throughout across all groups.

##### *(III) Computational model-derived learning parameters*

Learning parameters estimated from the computational model i.e. α (learning rate) and β (exploitation-exploration index or randomness) were used to ascertain (i) optimal functioning of algorithm, and (ii) consistency with performance deduced from observed choices. Surprisingly, α and β estimates were at upper and lower extremes, that is, concentrated at 0 or 1, for ≥ 50% of participants under both conditions (**Figure S8**). This was unequivocal evidence that the computational model was not functioning optimally and, thereby, not modelling observed choices accurately. No significant between-group differences were noted for both parameters under either condition. To establish a manual link to poor task comprehension in participants based on β distribution, I also visually examined motor performance (button-pressing activity) throughout the course of trials and indeed noted abnormal response patterns, e.g. absent button-pressing, or sustained button pressing, in a number of participants (N=5) (**Figure S9**).

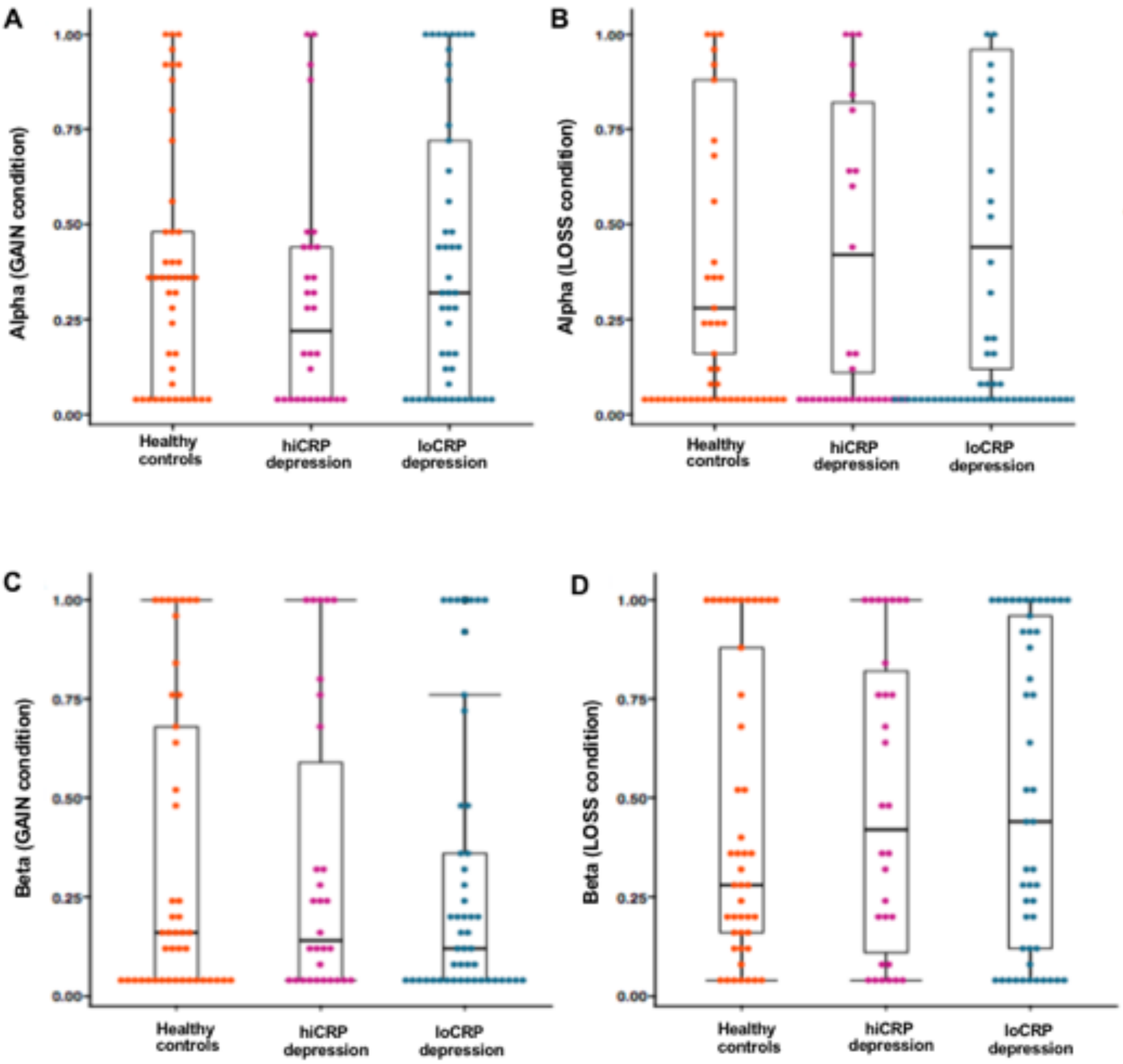


**Figure S8 Computational model parameters (A-B) α (learning rate) and (C-D) β (exploitation- exploration index).** The Q-learning algorithm was likely not modelling observed behaviour optimally, as parameters estimated were at extreme values for nearly 50% of participants per group.

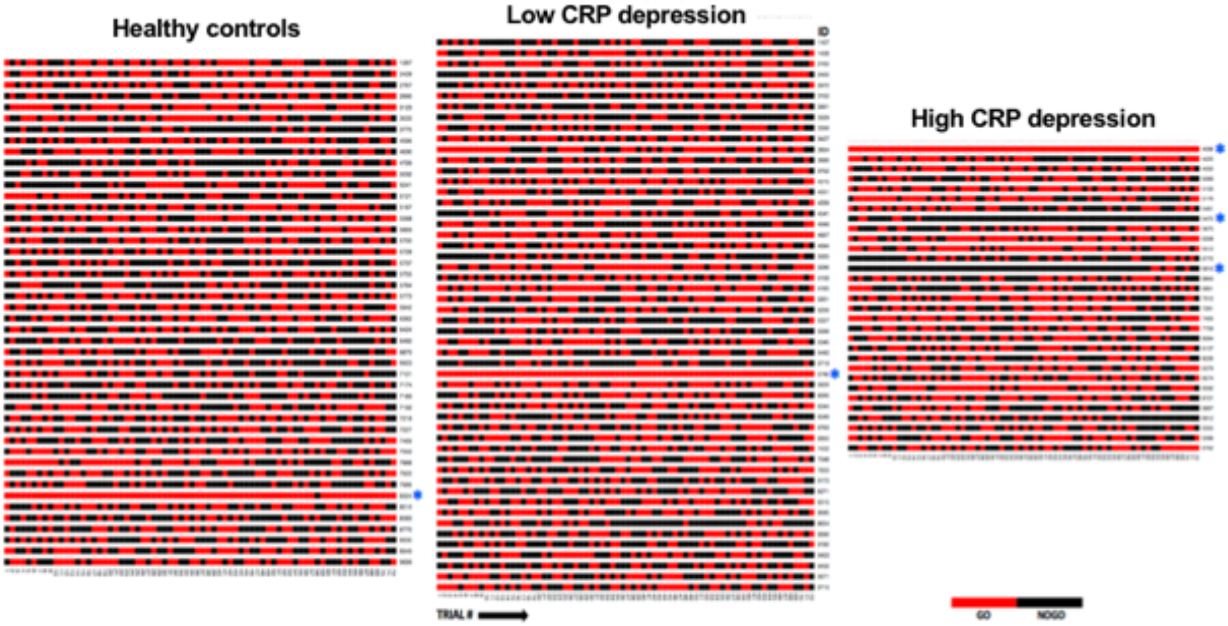


**Figure S9 Motor performance (button pressing) pattern across trials.** Participants denoted with blue asterisk were identified to have abnormal motor activity e.g. continuous pressing, or absent pressing across trials.

##### *(IV) Functional activation to Gain and Loss monetary cues*

To establish fMRI concordance of poor learning and suboptimal task comprehension in participants, I finally examined functional activation to ascertain if anticipation of monetary reward and punishment during presentation of stimuli pair reliably activated relevant brain regions. The GO > NO-GO contrast (choice or button-pressing event) was examined initially as a positive control to ensure GLM modelling were implemented accurately (**Figure S10**). As anticipated from poor quality data derived from task-related behavior, no significant within- group effects were noted for reward stimuli pair presentation (GAIN > LOOK contrast) (**Figure S11**) or punishment stimuli pair presentation (LOSS > LOOK contrast) (**Figure S12**) across all groups. Predictably, no significant between-group effects were noted for either stimuli pair contrasts.

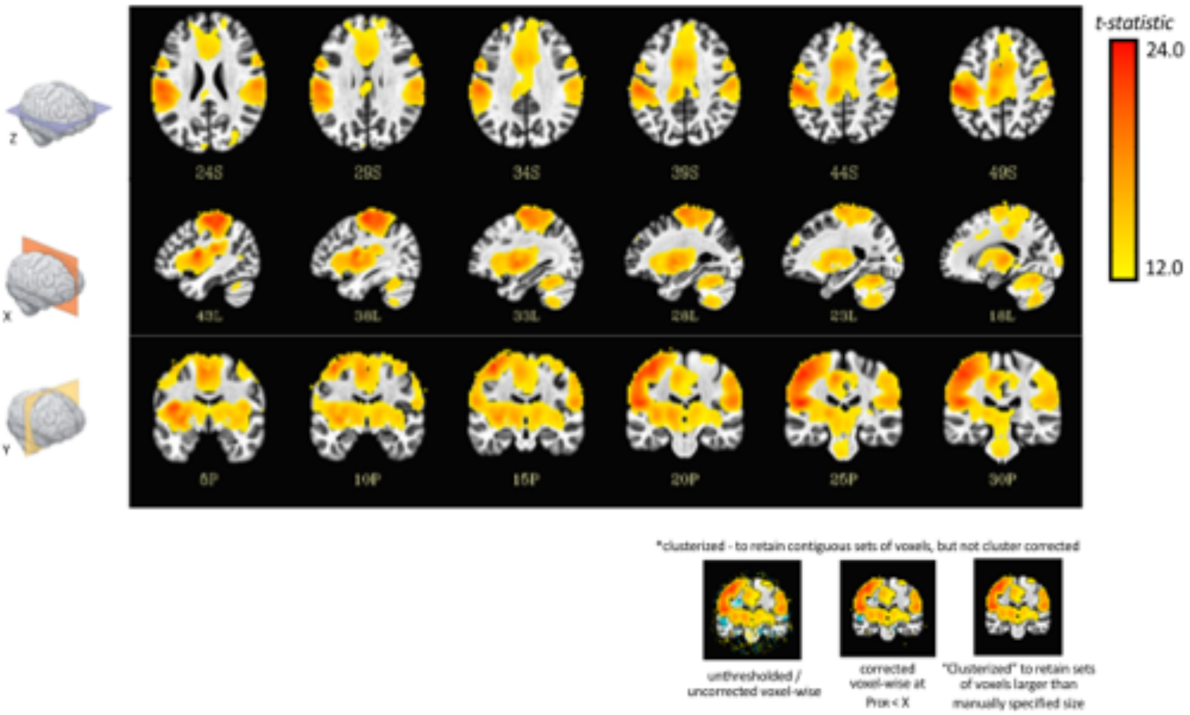


**Figure S10 Neural activation to button pressing (GO > NO-GO contrast).** Prominent contralateral activation of motor cortices and downstream motor pathway during button-pressing or choice event. All activation maps are depicted in MNI space in neurological convention (left side of the brain on left side of the picture). The color bar depicts t-values of local maxima peak activation.

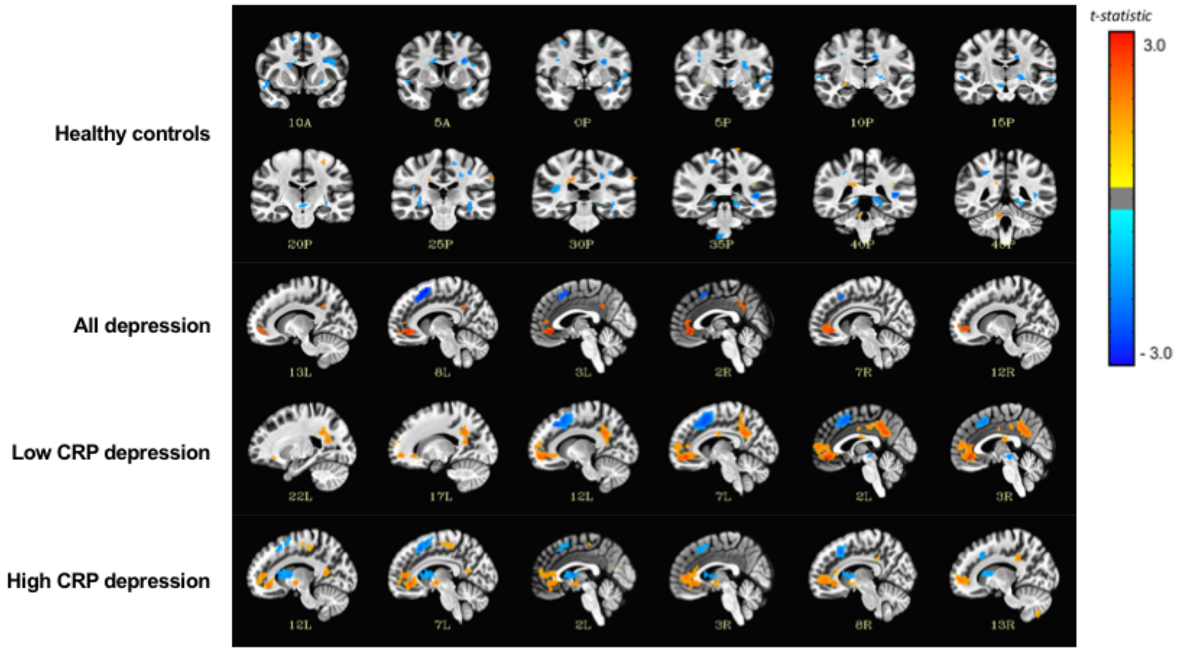


**Figure S11 Functional activation to presentation of Gain stimuli (GAIN > LOOK contrast).** No significant within-group activation upon presentation of stimuli pair under Gain condition. All activation maps are depicted in MNI space in neurological convention (left side of the brain on left side of the picture). The color bar depicts t-values of local maxima peak activation.

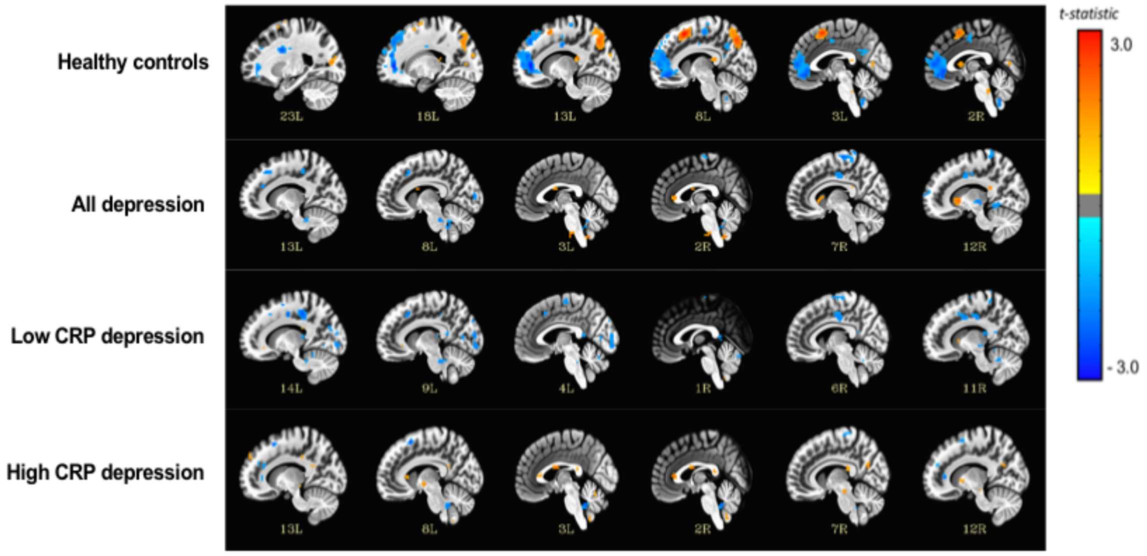


**Figure S12 Functional activation to presentation of Loss stimuli (LOSS > LOOK contrast).** No significant within-group activation upon presentation of stimuli pair under Loss condition. All activation maps are depicted in MNI space in neurological convention (left side of the brain on left side of the picture). The color bar depicts t-values of local maxima peak activation.

*Task paradigm QC outcome*

In sum, it was evident after QC that task comprehension and quality of the behavioral data was suboptimal, with potential repercussions on suitability of task-fMRI data for relevant hypothesis testing. This led to revision of the final analytic plan for task-fMRI paradigm, whereby the computational model-derived prediction error (δ) was excluded from inclusion as an amplitude modulator in general linear modelling (GLM) of first-level fMRI timeseries analysis, as had performed in other similar studies of probabilistic reinforcement learning (1,3). Additionally, anticipation or stimuli presentation events were not examined as part of hypothesis testing. The five participants (N=1 HC, N=1 low CRP depression, N=3 high CRP depression) noted to have abnormal button-pressing activity were excluded from subsequent analyses as this was an unequivocal mark of limited task comprehension overall and may bias statistical results.
